# Respiratory ciliary abnormalities in heterotaxy beyond primary ciliary dyskinesia: functional and ultrastructural characterisation

**DOI:** 10.64898/2026.03.17.26348660

**Authors:** Laura Venditto, Mathieu Bottier, Ranjit Rai, Rhiannon Mclellan, Georgina Bailey, Emily Howieson, Mellisa Dixon, Samantha J Irving, Deborah J Morris-Rosendahl, Amelia Shoemark, Claire Hogg, Thomas Burgoyne

**Affiliations:** Department of Surgery, Dentistry, Paediatrics and Gynaecology, University of Verona, Verona, Italy; Paediatric Respiratory Medicine, Primary Ciliary Dyskinesia Centre, Royal Brompton Hospital, Guy’s and St Thomas NHS Foundation Trust, London, United Kingdom; National Heart and Lung Institute, Imperial College London, London, United Kingdom; Clinical Genetics and Genomics Laboratory, Royal Brompton Hospital, Guy’s and St. Thomas NHS Foundation Trust, London, United Kingdom; University of Dundee, Dundee, United Kingdom; Institute of Ophthalmology, University College London, London, United Kingdom

**Author notes:** Shared first authorship (Laura Venditto, Mathieu Bottier). **Corresponding Author** Dr Thomas Burgoyne UCL Institute of Ophthalmology, University College London, 11-43 Bath St, London EC1V 9EL (United Kingdom).

**Keywords:** Airway epithelium, rare lung disease, heterotaxy, congenital heart disease

## Abstract

**Background:** Heterotaxy (HTX) is characterised by abnormal left-right arrangement of the internal organs and is frequently associated with complex congenital heart disease (CHD). HTX is enriched among patients with primary ciliary dyskinesia (PCD), due to impaired function of embryonic nodal cilia involved in left-right patterning. However, a subset of patients with HTX presents with mild or absent respiratory phenotypes and normal respiratory ciliary function, and the mechanisms underlying situs abnormalities in these non-PCD patients remain unclear. We aimed to compare the clinical history, respiratory ciliary structure, and ciliary function of patients with HTX with and without PCD.

**Methods:** Retrospective diagnostic data were analysed from 73 subjects with HTX referred to the Royal Brompton Hospital PCD Diagnostic Service between 1997 and 2024. Of these, 30 were diagnosed with PCD (HTX-PCD) and 43 did not meet the diagnostic criteria for PCD (HTX-only). Data included clinical history, high-speed video-microscopy (HSVM), transmission electron microscopy (TEM) of ciliary ultrastructure, PCD genotype and cardiac and thoracic/abdominal situs imaging.

**Results:** Atrioventricular discordance (*p* = 0.0102) and midline liver position (*p* = 0.0377) were more frequent in the HTX-only group. TEM analysis identified extra microtubules in 24.4% of HTX-only subjects and microtubular disorganisation in 24.4%. Review of TEM data from 2,823 individuals referred for PCD assessment demonstrated that these ultrastructural abnormalities were associated with an increased prevalence of CHD (*p* = 0.04). Quantitative HSVM analysis demonstrated preserved or increased ciliary beat amplitude in HTX-only compared with HTX-PCD patients.

**Discussion:** HTX without PCD is associated with distinct laterality phenotypes and subtle respiratory ciliary abnormalities below current PCD diagnostic thresholds. These findings suggest that mild defects affecting ciliary structure and function may contribute to abnormal left-right patterning without causing classical PCD. Larger multicentre genetic and phenotypic studies are required to further define the mechanisms underlying HTX.

## Introduction

The word heterotaxy (HTX) is derived from Greek, in which *heteros*-means “other than” and *taxis*-“arrangement”. It describes a pattern of anatomical organisation of thoracic and abdominal organs that differs from situs solitus and situs inversus totalis (1). Patients can present an unusual degree of symmetry of the thoracic and abdominal organs, along with the atrial appendages of the heart, including a wide variety of complex cardiac lesions. Patients with HTX can be stratified into the subsets: asplenia syndrome (absence of the spleen), HTX with isomerism of the right atrial appendages, and polysplenia syndrome or HTX with isomerism of the left atrial appendages (1). Cardiac anatomy in HTX patients varies from normal or nearly normal biventricular circulation to complex single ventricle congenital heart defects (CHD). Certain CHDs are more common in patients who have right atrial isomerism rather than left atrial isomerism (2).

While the prevalence of HTX in the general population has been reported as 1 in 10,000 total births (3), it is much higher in Primary Ciliary Dyskinesia (PCD) patientreported between 6.3% (4) and 12.1% (5,6). This is believed to be due to dysfunction in motility of embryonic nodal cilia that are required to establish normal left-right asymmetry (7). A number of studies that assessed CHD patients with HTX, have shown a link between abnormal respiratory cilia motility, reduced nasal nitric oxide and a respiratory disease phenotype (8–13). Some studies have provided evidence that patients with HTX could have ciliary dysmotility, which could have an impact on their morbidity, especially after cardiac surgery (10,14–16). Comparing studies can be difficult as the methods used to assess cilia function vary along with the definitions of HTX.

Despite previous studies, it remains unclear why some subjects with HTX who do not have PCD, and who appear to have normal motile respiratory ciliary function, develop abnormal situs and exhibit mild respiratory symptoms. In this study, we compared retrospective diagnostic data from patients with HTX referred for PCD diagnostic testing, including reanalysis of high-speed video-microscopy (HSVM) videos and transmission electron microscopy (TEM) images to comprehensively assess respiratory ciliary motility and ultrastructure. We hypothesised that a subset of patients with HTX without PCD harbour subtle abnormalities in respiratory ciliary function or ultrastructure that fall below the current diagnostic thresholds for PCD but nevertheless reflect underlying defects in motile cilia that contribute to abnormal left–right patterning during embryonic development and to mild respiratory disease.

## Materials and methods

### Study design

Retrospective data from subjects who were referred for PCD diagnostic testing at the Royal Brompton Hospital between 1997 and 2024 were included in this study (Guy’s and St Thomas NHS Foundation Trust Electronic Record Research Interface (GERRI) under IRAS ID: 257283 and Rec Reference: 20/EM/0112).

### Study cohort

Seventy-three subjects were identified with HTX alongside twenty-eight control individuals that were included in the study that had been referred for PCD diagnostic assessment who did not have HTX and were ultimately classified as not having PCD based on normal diagnostic test results (defined as non-HTX, non-PCD). A TEM secondary study included reports from 2,823 individuals referred for diagnostic testing for PCD.

### PCD diagnosis

PCD diagnosis was established at the Royal Brompton Hospital in accordance to the European Respiratory Society (ERS) guidelines at the time of diagnosis (17–19). Diagnostic data included nasal Nitric Oxide (nNO) measurements when feasible, HSVM analysis reports (20), TEM ultrastructure reports (21) and genetic testing where available. Individuals with low clinical suspicion and normal nNO and HSVM findings were not routinely referred for genetic testing. Individuals with a positive or highly suggestive diagnosis of PCD underwent testing using the Respiratory Ciliopathies including non-CF Bronchiectasis panel (Genomics England R189), whereas those with heterotaxy or other laterality abnormalities but lower suspicion of PCD were referred for the Laterality Disorders and Isomerism panel (Genomics England R139), which also includes PCD-associated genes implicated in laterality defects.

### HTX and Organ Defect Classification

Medical reports were reviewed to determine cardiac and abdominal situs using echocardiography, abdomen ultrasound and CT scan results. We defined HTX as an abnormal situs that is neither *situs solitus* nor *situs inversus*. This included those patients with an abnormal or discordant situs in either pulmonary, gastrointestinal, splenic, hepatobiliary and/or cardiovascular system. This includes polysplenia, asplenia, pulmonary or bronchial isomerism, midline or left-sided liver, malrotation, midline or right-sided stomach. CHD patients that only have situs related cardiovascular anomalies were also included, such as dextrocardia or mesocardia, interrupted inferior vena cava, atrial situs inversus, ambiguous or isomerism, atriovascular discordance, superior/inferior ventricles, and ventricular-arterial discordance.

### Ciliary Beat Pattern Analysis

Using archived retrospective HSVM videos (previously collected on a Leica DM-LB upright microscope fitted with a Troubleshooter TS-5 Fastec imaging camera), high-speed video-microscopy analysis (HSVA) was undertaken using in-house software developed using Matlab R2023A based on a previously published method (22–24). By selecting a cilium in each video that could be tracked during a complete beat cycle, parameters including ciliary beat frequency (CBF), cilium length, ciliary beating angle, ciliary beating amplitude, and amplitude per second (defined as amplitude x CBF) were obtained. The videos from all subjects were blinded before assessment to reduce bias.

### Statistical Analysis

Frequencies and percentages were calculated for categorical variables, and median and range for continuous variables. Fisher exact test was used to test differences in categorical variables. Mann-Whitney and Kruskal-Wallis tests were used to test for differences in continuous variables. P-values less than 0.05 were considered statistically significant. Analyses were performed using GraphPad Prism version 11.0.2 for Windows (GraphPad Software, Boston, MA, USA) and missing patient data was excluded from the analysis as indicated in the results tables.

### Patient and Public Involvement

This was a retrospective cohort study, and no patient or public partners were involved in the design, conduct or dissemination of this research.

## Results

From the seventy-three HTX subjects, a diagnosis of PCD was confirmed or considered highly likely in 30/73 patients (41.1%) by molecular genetics, TEM or both (defined as HTX-PCD) and excluded or considered unlikely in 43/73 (defined as HTX-only). Grouping HTX patients according to their PCD diagnosis enabled us to identify key clinical similarities and differences between patients with and without PCD.

### Genetic variants

In the HTX-PCD group, 24/30 (80.0%) subjects underwent genetic testing (Genomics England R189). Genetic reports were reviewed by specialist clinical geneticists, and only pathogenic or likely pathogenic variants reported as clinically diagnostic of PCD were included in this study. In total, 21 of 24 subjects (87.5%) had a pathogenic or likely pathogenic variants identified in a genotype consistent with the established inheritance pattern of the respective PCD-associated gene. No pathogenic or likely pathogenic variants were identified in the remaining three subjects; however, PCD was confirmed in these individuals by characteristic class 1 TEM defects. The distribution of disease-causing genes and their corresponding functional defects is shown in Figure 1. Consistent with previous international studies, pathogenic variants were most frequently identified in *DNAH5* and *DNAH11*, which encode outer dynein arm proteins. As expected, no pathogenic variants were identified in genes encoding proteins absent from embryonic nodal cilia, such as those exclusively associated with the central apparatus or radial spokes.

**Figure 1.**
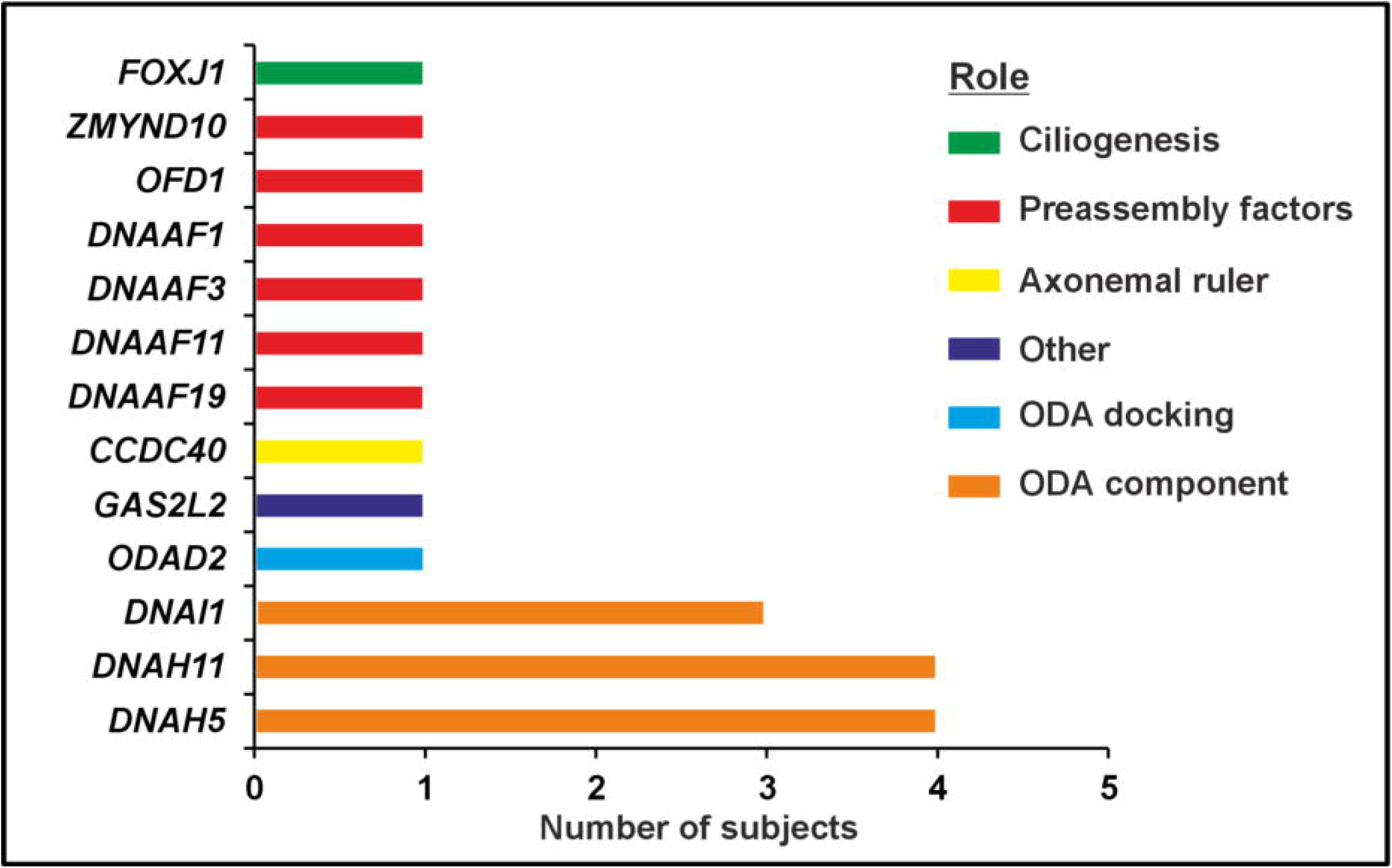
Genes identified with pathogenic variants in HTX patients with PCD. The relative function of the encode protein from these genes is shown. Preassembly factors encode cytoplasmic protein involved in the assembly of inner and outer dynein arms (ODA).

In the HTX-only group, genetic testing (Genomics England R139) was performed in 8/43 (18.6%) subjects. Only one subject received a genetic diagnosis, being homozygous for a pathogenic variant in the *MNS1* gene. *MNS1* is associated with autosomal recessive heterotaxy with male infertility and has been linked to a mild respiratory phenotype but is currently not associated with PCD (25).

### PCD related clinical features

HTX patients with and without PCD were referred for investigation at a similar age (median 1.25 and 0.75 years old, respectively). When examining respiratory symptoms classically associated with PCD, these were present in 27/30 patients (90.0%) in the HTX-PCD group compared with 21/43 patients (48.8%) in the HTX-only group (see Table 1). Respiratory symptoms were a reason for referral for PCD diagnosis; despite this, there was a statistically significant difference in persisting wet cough, rhinitis, and presence of respiratory distress at birth, which were more frequent in the HTX-PCD group. There was no difference in rate of recurrent chest infections between groups.

**Table 1.**
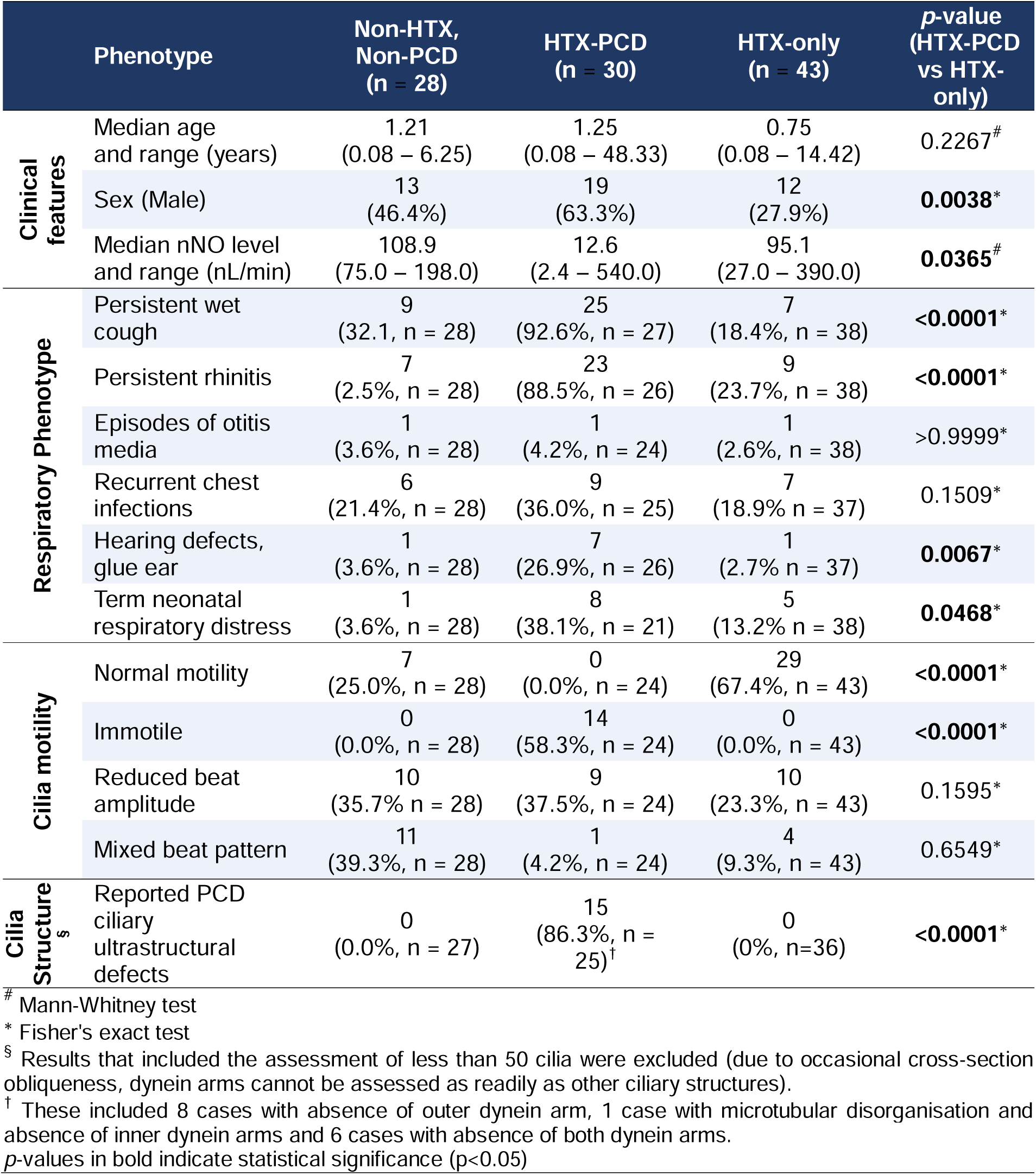
PCD-related respiratory phenotype and diagnostic results at the time of the referral. . Percentages shown are based on number of patients that had available reports, which are indicated in the brackets.

The original diagnostic reports were reviewed to extract the measured level of nNO, detailed descriptions of ciliary motility on HSVM and ciliary ultrastructural defects on TEM, which are summarised in Table 1. Supplemental table 1 highlights the difference between the control group and both HTX groups.

There were limited nNO measurements because many patients were too young to undergo nNO measurement at the time of referral. nNO measurements were available for 12 of 30 patients (40 %) in the HTX-PCD group, 5 of 43 patients (11.62%) in the HTX-only group, and 3 of 28 control subjects (10.71%). In the HTX-PCD group, the average nNO value was significantly lower (12.6 nl/min) compared to the HTX-only group (95.1 nl/min, *p* = 0.0365). HSVM reports were available for 24 of 30 subjects (80.0%) in the HTX-PCD group, 42 of 43 subjects (97.7%) in the HTX-only group, and all 28 control subjects (100.0%). The HTX-only group included eight repeat nasal brushings. TEM reports were available for 26 of 30 subjects in the HTX-PCD group, 36 of 43 subjects (83.7%) in the HTX-only group, and 27 of 28 control subjects (96.4%). Only the HTX-PCD group reported ultrastructural defect (86.3% of samples): 8 cases with outer dynein arm absence, 1 case with microtubular disorganisation and inner dynein arms absence and 6 cases with absence of both dynein arms. Due to the high prevalence of dynein arms defects in the HTX-PCD group, it was not surprising that there were many immotile cilia and other motility abnormalities.

### Situs abnormalities

The spectrum of situs abnormalities in HTX patients was determined from the collected clinical data. Table 2 summarises the organ situs abnormalities. These analyses exclude patients with indeterminate situs or unavailable imaging. The non-PCD/non-HTX control group is not included, as no situs, cardiac, or vascular abnormalities were identified in these individuals. Among patients with available imaging, 14 of 28 (50.0%) in the HTX-PCD group and 12 of 42 (28.6%) in the HTX-only group had abnormal cardiac position, with no significant difference between the groups (*p* = 0.25). This included one patient in the HTX-only group with mesocardia. Interestingly, a midline liver was significantly more common in the HTX-only group (*p* = 0.0377).

**Table 2.**
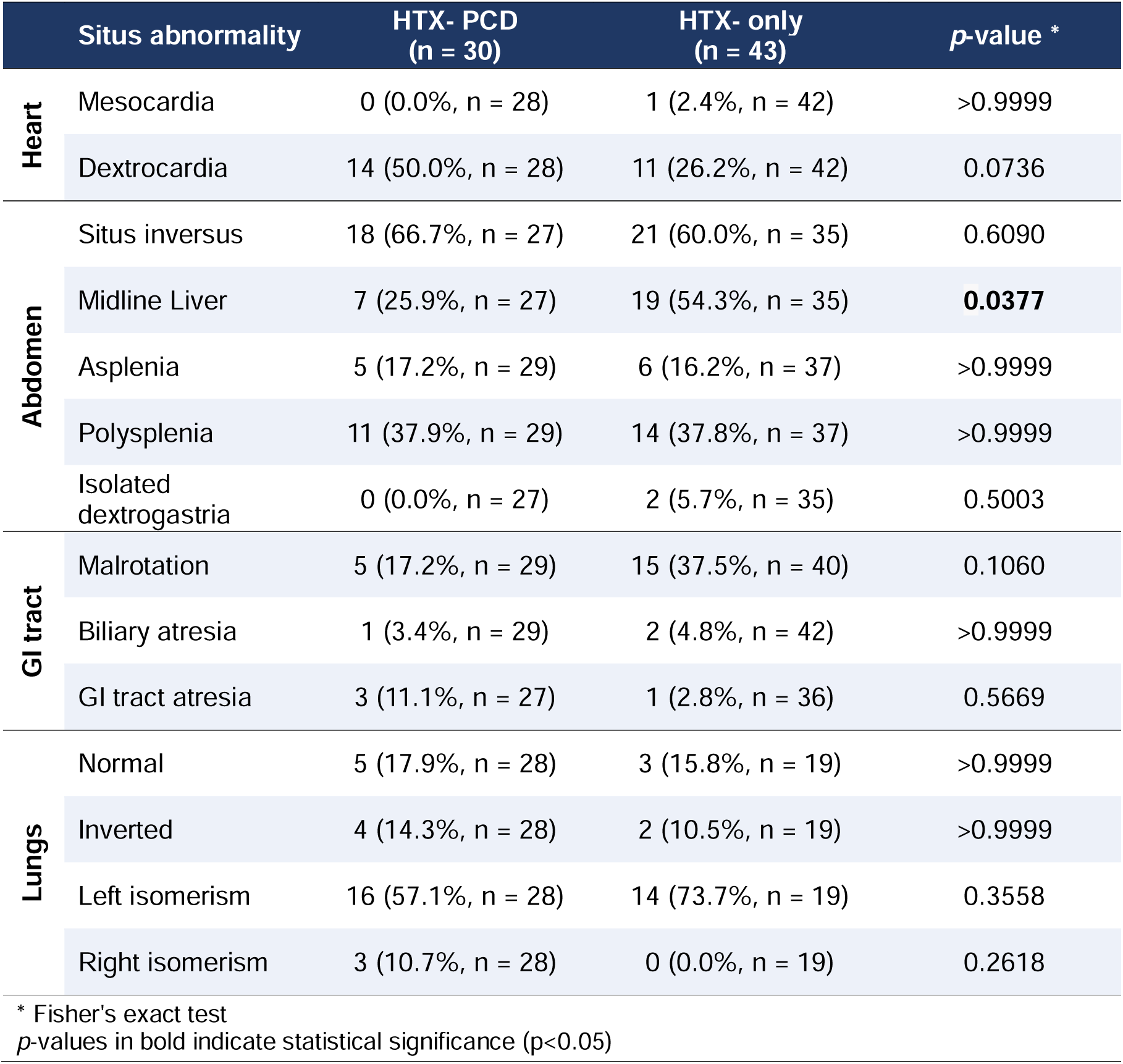
A comparison of organ situs abnormalities in subject with HTW with or without PCD.

|  | Situs abnormality | HTX- PCD<br>(n = 30) | HTX- only<br>(n = 43) | p-value * |
| --- | --- | --- | --- | --- |
| Heart | Mesocardia | 0 (0.0%, n = 28) | 1 (2.4%, n = 42) | >0.9999 |
|  | Dextrocardia | 14 (50.0%, n = 28) | 11 (26.2%, n = 42) | 0.0736 |
| Abdomen | Situs inversus | 18 (66.7%, n = 27) | 21 (60.0%, n = 35) | 0.6090 |
|  | Midline Liver | 7 (25.9%, n = 27) | 19 (54.3%, n = 35) | **0.0377** |
|  | Asplenia | 5 (17.2%, n = 29) | 6 (16.2%, n = 37) | >0.9999 |
|  | Polysplenia | 11 (37.9%, n = 29) | 14 (37.8%, n = 37) | >0.9999 |
|  | Isolated dextrogastria | 0 (0.0%, n = 27) | 2 (5.7%, n = 35) | 0.5003 |
| GI tract | Malrotation | 5 (17.2%, n = 29) | 15 (37.5%, n = 40) | 0.1060 |
|  | Biliary atresia | 1 (3.4%, n = 29) | 2 (4.8%, n = 42) | >0.9999 |
|  | GI tract atresia | 3 (11.1%, n = 27) | 1 (2.8%, n = 36) | 0.5669 |
| Lungs | Normal | 5 (17.9%, n = 28) | 3 (15.8%, n = 19) | >0.9999 |
|  | Inverted | 4 (14.3%, n = 28) | 2 (10.5%, n = 19) | >0.9999 |
|  | Left isomerism | 16 (57.1%, n = 28) | 14 (73.7%, n = 19) | 0.3558 |
|  | Right isomerism | 3 (10.7%, n = 28) | 0 (0.0%, n = 19) | 0.2618 |

\* Fisher's exact test
p-values in bold indicate statistical significance (p<0.05)

The cardiovascular abnormalities identified in the HTX cohort are summarised in Table 3. No cardiac or vascular defect was more prevalent in either group, although atrioventricular discordance was significantly more common in the HTX-only group (*p* = 0.0102).

**Table 3.**
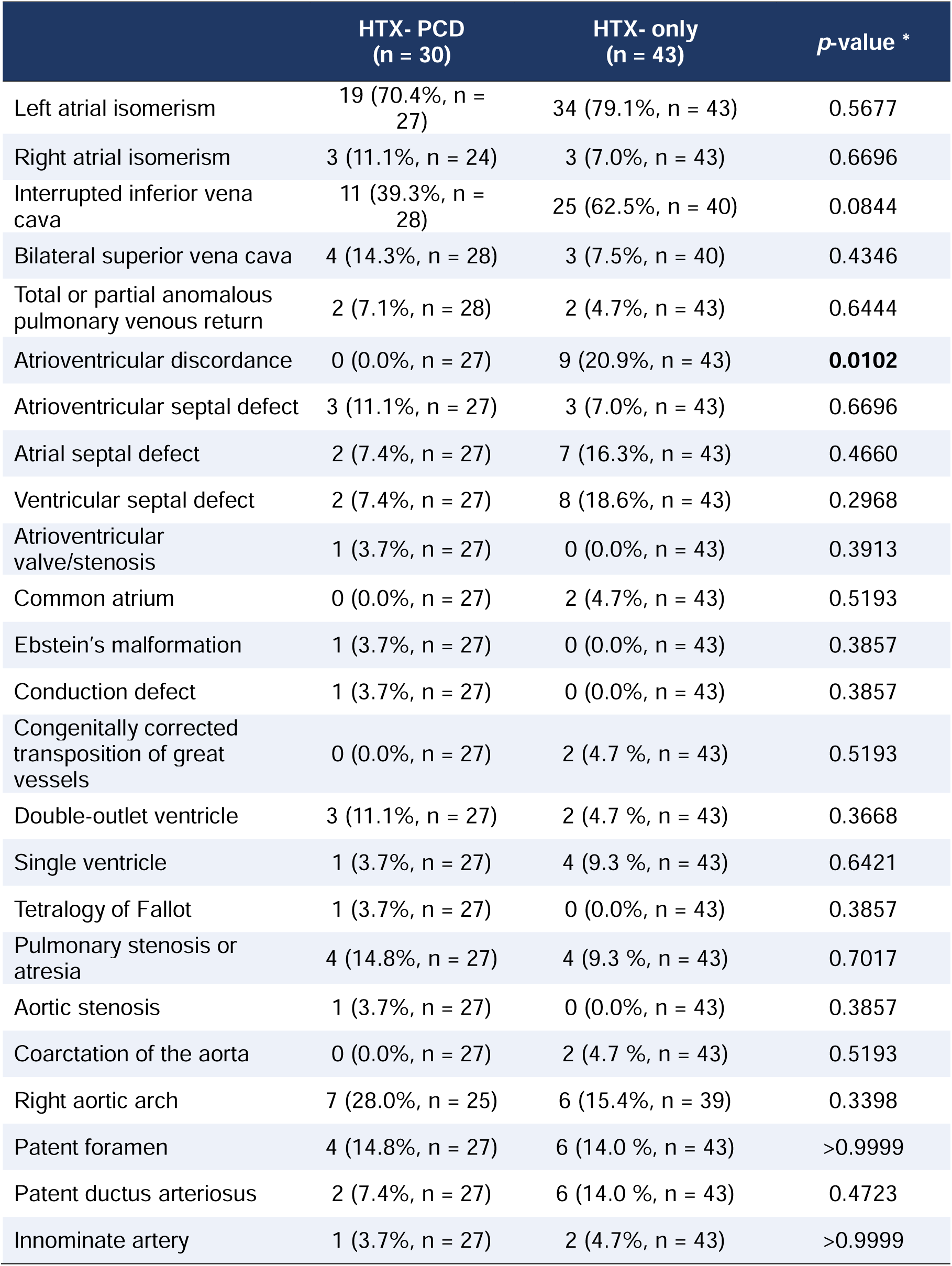
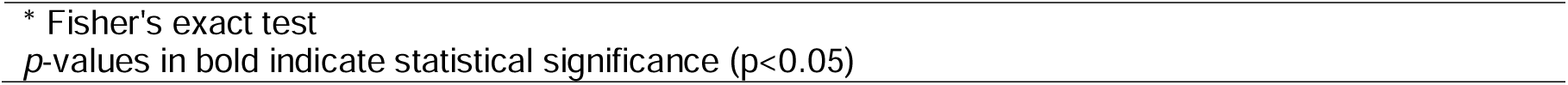
Types of cardiovascular anomalies found in subjects with HTX with or without PCD.

|  | HTX- PCD<br>(n = 30) | HTX- only<br>(n = 43) | p-value * |
| --- | --- | --- | --- |
| Left atrial isomerism | 19 (70.4%, n = 27) | 34 (79.1%, n = 43) | 0.5677 |
| Right atrial isomerism | 3 (11.1%, n = 24) | 3 (7.0%, n = 43) | 0.6696 |
| Interrupted inferior vena cava | 11 (39.3%, n = 28) | 25 (62.5%, n = 40) | 0.0844 |
| Bilateral superior vena cava | 4 (14.3%, n = 28) | 3 (7.5%, n = 40) | 0.4346 |
| Total or partial anomalous pulmonary venous return | 2 (7.1%, n = 28) | 2 (4.7%, n = 43) | 0.6444 |
| Atrioventricular discordance | 0 (0.0%, n = 27) | 9 (20.9%, n = 43) | **0.0102** |
| Atrioventricular septal defect | 3 (11.1%, n = 27) | 3 (7.0%, n = 43) | 0.6696 |
| Atrial septal defect | 2 (7.4%, n = 27) | 7 (16.3%, n = 43) | 0.4660 |
| Ventricular septal defect | 2 (7.4%, n = 27) | 8 (18.6%, n = 43) | 0.2968 |
| Atrioventricular valve/stenosis | 1 (3.7%, n = 27) | 0 (0.0%, n = 43) | 0.3913 |
| Common atrium | 0 (0.0%, n = 27) | 2 (4.7%, n = 43) | 0.5193 |
| Ebstein's malformation | 1 (3.7%, n = 27) | 0 (0.0%, n = 43) | 0.3857 |
| Conduction defect | 1 (3.7%, n = 27) | 0 (0.0%, n = 43) | 0.3857 |
| Congenitally corrected transposition of great vessels | 0 (0.0%, n = 27) | 2 (4.7 %, n = 43) | 0.5193 |
| Double-outlet ventricle | 3 (11.1%, n = 27) | 2 (4.7 %, n = 43) | 0.3668 |
| Single ventricle | 1 (3.7%, n = 27) | 4 (9.3 %, n = 43) | 0.6421 |
| Tetralogy of Fallot | 1 (3.7%, n = 27) | 0 (0.0%, n = 43) | 0.3857 |
| Pulmonary stenosis or atresia | 4 (14.8%, n = 27) | 4 (9.3 %, n = 43) | 0.7017 |
| Aortic stenosis | 1 (3.7%, n = 27) | 0 (0.0%, n = 43) | 0.3857 |
| Coarctation of the aorta | 0 (0.0%, n = 27) | 2 (4.7 %, n = 43) | 0.5193 |
| Right aortic arch | 7 (28.0%, n = 25) | 6 (15.4%, n = 39) | 0.3398 |
| Patent foramen | 4 (14.8%, n = 27) | 6 (14.0 %, n = 43) | >0.9999 |
| Patent ductus arteriosus | 2 (7.4%, n = 27) | 6 (14.0 %, n = 43) | 0.4723 |
| Innominate artery | 1 (3.7%, n = 27) | 2 (4.7%, n = 43) | >0.9999 |

\* Fisher's exact test
*p*-values in bold indicate statistical significance ( $p < 0.05$ )

### Review of ciliary ultrastructure assessment

In the HTX-PCD, the most common ultrastructural defects were class 1 absence of outer dynein arms (32.0%) or class 1 absence of both outer and inner dynein arms (24.0%), aligning with pathogenic variants in dynein structural and assembly genes (Figure 1). One patient had a likely nexin-dynein regulatory complex defect (class 2 absence of inner dynein arms), while 36.0% showed normal ciliary ultrastructure.

For the HTX-only group, TEM was conducted in all, except two samples which were inadequate for analysis. As expected, no PCD related ultrastructural defects were found. Instead, secondary abnormalities, microtubular disorganisation and extra microtubules, often linked to infection or inflammation were examined (Figure 2). Among HTX-only samples, 10 of 41 (24.4%) showed microtubular disorganisation affecting >5% of cilia, and 10 of 41 (24.4%) showed extra microtubules in >5% of cilia.

**Figure 2.**
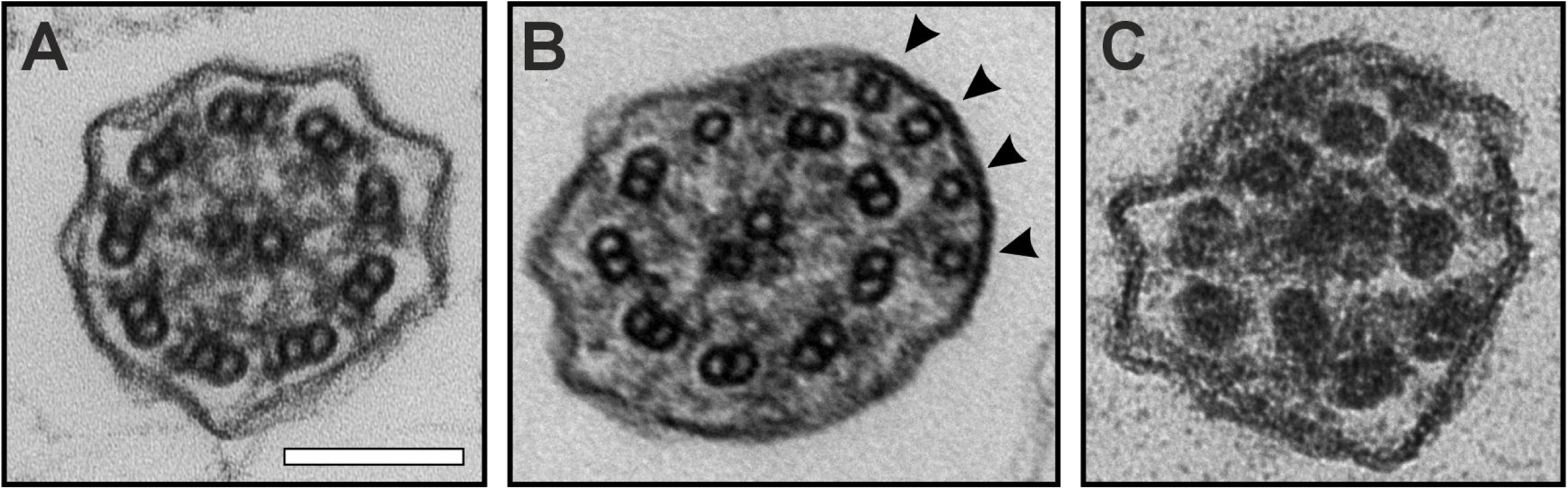
Representative transmission electron microscopy images of respiratory ciliary ultrastructure. (A) Normal ciliary axoneme showing the typical 9+2 arrangement, with nine peripheral microtubule doublets surrounding two central microtubules and intact outer and inner dynein arms. (B) Ciliary axoneme with extra microtubules (arrowheads). (C) Ciliary axoneme exhibiting microtubular disorganization. Scale bar: 100 nm.

To investigate the association between HTX and these ultrastructural defects, TEM data from 2,823 individuals referred for PCD diagnostic assessment between 1997 and 2024 were retrospectively reviewed. Subjects with ≥10% of cilia affected by either defect were cross-referenced for the presence of cardiac and situs abnormalities. Only individuals with ≥50 analysed ciliary cross-sections were included in the analysis. Diagnostic reports were reviewed for PCD, CHD, dextrocardia, and abnormal situs (Table 4). Non-PCD subjects with extra microtubules had higher rates of CHD and dextrocardia compared to controls, while those with microtubular disorganisation were more likely to have CHD. Overall, microtubular disorganisation was associated with a greater likelihood of abdominal situs defects.

**Table 4.** Distribution of cardiac defects, dextrocardia and situs abnormalities in subjects with disarrangement or extra microtubule ciliary defects.

| Type of defect | Patient group | Control*<br>(non-PCD n = 90,<br>PCD n = 38) | Microtubular<br>disorganisation<sup>†</sup> (non-PCD n = 40, PCD n =<br>59) | Extra microtubules<sup>†</sup><br>(non-PCD n = 154,<br>PCD n = 35) |
| --- | --- | --- | --- | --- |
| CHD | All | 17 (13.28%) | 27 (27.27%)<br>$p = \mathbf{0.0082}$ | 42 (22.22%)<br>$p = 0.0555$ |
| | PCD | 13 (34.21%) | 20 (33.90%)<br>$p > 0.999$ | 20 (57.14%)<br>$p = 0.0619$ |
| | Non-PCD | 4 (4.44%) | 7 (17.50%)<br>$p = \mathbf{0.0342}$ | 22 (14.29%)<br>$p = \mathbf{0.0175}$ |
| Dextro-cardia | All | 13 (10.16%) | 17 (17.17%)<br>$p = 0.1659$ | 32 (16.93%)<br>$p = 0.1022$ |
| | PCD | 11 (28.95%) | 15 (25.42%)<br>$p = 0.8151$ | 17 (48.57%)<br>$p = 0.0978$ |
| | Non-PCD | 2 (2.22%) | 2 (5.00%)<br>$p = 0.5863$ | 15 (9.74%)<br>$p = \mathbf{0.0346}$ |
| Abdominal<br>Situs defect | All | 18 (14.01%) | 27 (27.27%)<br>$p = \mathbf{0.0183}$ | 33 (17.46%)<br>$p = 0.4409$ |
| | PCD | 13 (34.21%) | 24 (40.58%)<br>$p = 0.6689$ | 18 (51.43%)<br>$p = 0.1606$ |
| | Non-PCD | 5 (5.56%) | 3 (7.50%)<br>$p = 0.7010$ | 15 (9.74%)<br>$p = 0.3354$ |

\* Patients samples that had no cilia with a microtubular disorganisation or extra microtubule defects from the 2,823 patients referred for diagnostic testing between 1997 – 2024.
<sup>†</sup> Patients with $\geq 10\%$ of cilia affected by the named defect out of 2,823 patients referred for PCD diagnostics (1997 – 2024).
$p$ -values calculated by Fisher's exact test by comparing to the control group.
$p$ -values in bold indicate statistical significance ( $p < 0.05$ )

### Quantitative Analysis of the Ciliary Beat Pattern

Detailed quantitative HSVA was applied to videos from 58 HTX subjects, including 23 with PCD and 35 without a PCD diagnosis, to provide a more comprehensive assessment of ciliary parameters than the original diagnostic evaluations (n = 512 videos). 14 subjects in the HTX-PCD (60.9%) with immotile cilia were excluded from the analysis due to their extreme motility phenotype. For comparison, videos from 28 control subjects (non-HTX, non-PCD) were analysed (n = 274 videos). The comparison between control, HTX-PCD and HTX-only subjects is shown in Figure 3 and Supplementary Figure 1. A significant higher beating amplitude was found when comparing the HTX-only subjects against either the control or HTX-PCD subjects (*p* = 0.0258 and 0.0109 respectively).

**Figure 3.**
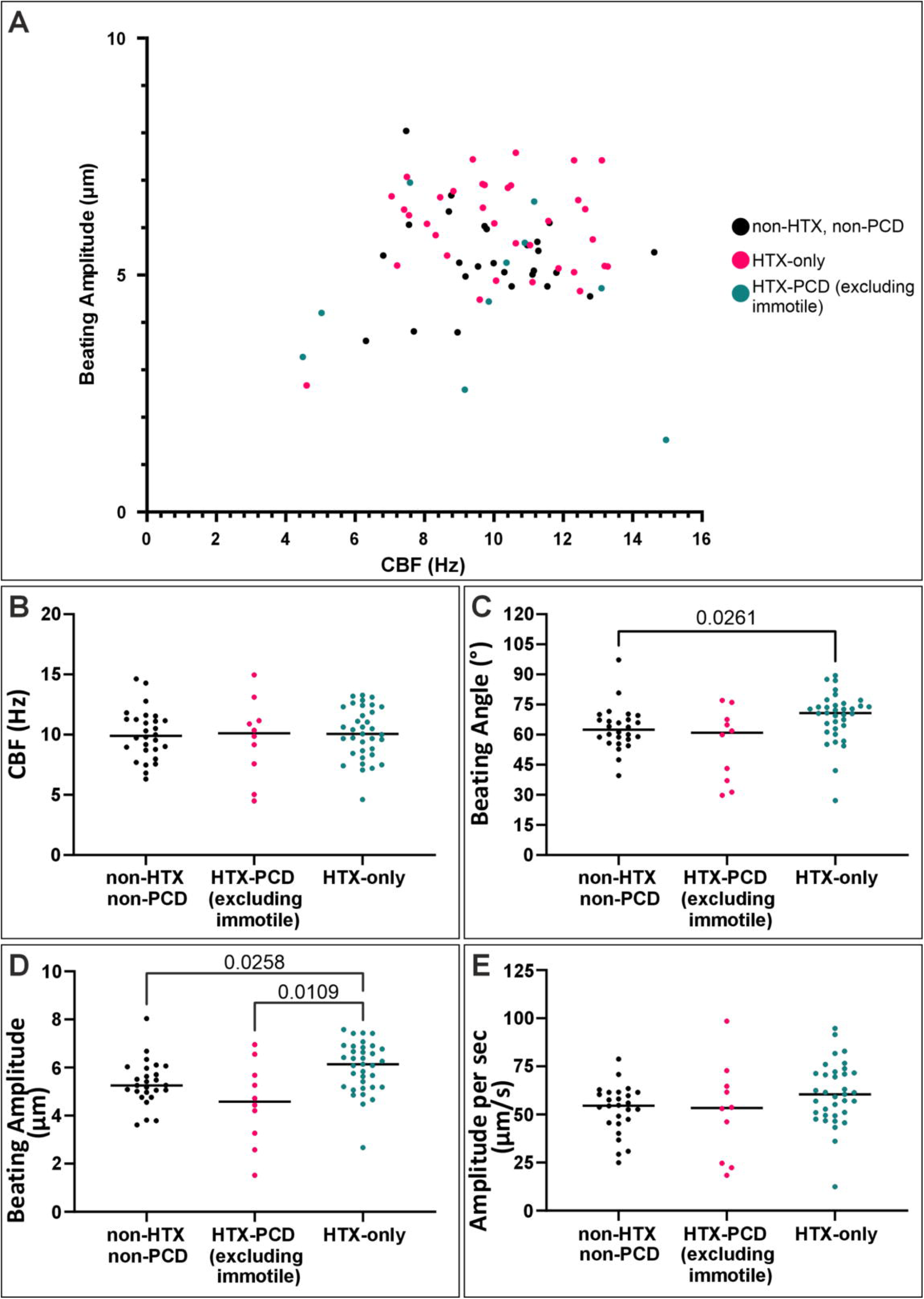
HTX-PCD subjects (excluding cases with static cilia) have reduced beating amplitude and angle compared to HTX-only subjects. (A) Comparison of the average cilia beat frequency (CBF) against average beating amplitude of cilia in controls (non-HTX non-PCD) and HTX patients with or without PCD. Average measures of (B) CBF, (C) beating angle, (D) beating amplitude and (E) amplitude per sec. (B – C) The line represents the median value. A significant difference in beating angle and beating amplitude was found between HTX patient with and without PCD. Statistical significance was determined by Kruskal-Wallis.

Focusing on the HTX-only group, we compared subgroups with ≥5% of cilia affected by microtubular disorganisation, ≥5% affected by extra microtubules, or <5% ultrastructural defects (No defect) on TEM (see Figure 4 and Supplementary Figure 2). Subjects with microtubular disorganisation and extra microtubules showed a significantly higher ciliary beat amplitude per second compared to controls (*p* = 0.0058 and 0.0452, respectively). They also had low epithelial disruption scores, indicating mainly normal strips of ciliated epithelial cells, with little evidence of infection (Supplementary Figure 2). These findings suggest that low-level ultrastructural abnormalities are associated with subtle impairments in ciliary function in patients with HTX who do not meet the diagnostic criteria for PCD.

**Figure 4.**
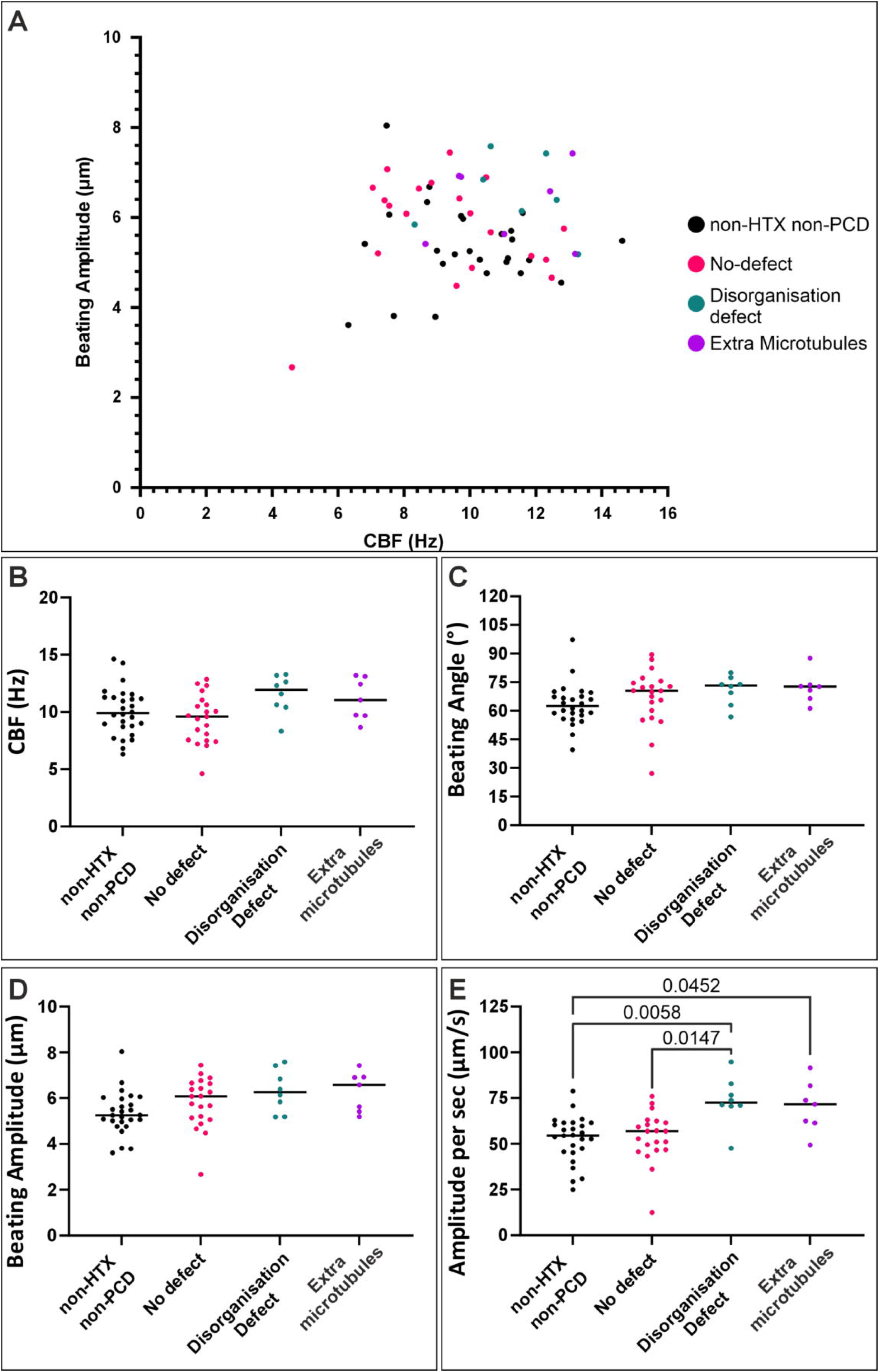
HTX-only subjects with >5% of cilia with a microtubular disorganization by electron microscopy had higher beating amplitude per second. (A) A comparison of the average cilia beat frequency (CBF) against average beating amplitude for controls (non-HTX non-PCD) and HTX-only subjects that had cilia with <5% of defects (No defect), >5% microtubular disorganization or >5% extra microtubules ciliary defects. Cases with microtubular disorganization and extra microtubule ciliary defects tended to have a higher (non-significant) CBF, beating angle and beating amplitude compared to cases without ultrastructural ciliary defects and controls. Average measures of (B) CBF, (C) beating angle, (D) beating amplitude and (E) amplitude per second. (B – E) The line represents the median value. There was a significant difference in the ciliary amplitude per second for cases with microtubular disorganization cilia compared to controls and those that had no defect. Statistical significance was determined by Kruskal-Wallis.

## Discussion

HTX represents a broad spectrum of disorders affecting left–right body patterning, with variable involvement of cardiac and other organ systems. The mechanisms underlying this phenotypic diversity remain incompletely understood. In this study, we compared diagnostic features of patients with HTX with and without PCD across the range of presentations encountered in clinical practice. These findings provide insights into the relationship between laterality defects, ciliary dysfunction, and respiratory phenotypes.

HTX and other situs abnormalities are enriched in patients with PCD due to shared protein composition between nodal and respiratory cilia. Defects in nodal cilia motility can disrupt embryonic left–right patterning, resulting in situs abnormalities (7). In contrast, the mechanisms underlying HTX in patients without PCD are more diverse. More than 40 genes associated with laterality defects but not PCD have been identified, including *LEFTY1*, *LEFTY2*, *NODAL*, and *PKD2* (26). These genes are involved in pathways downstream of nodal flow, suggesting that laterality defects can arise despite functional cilia. This may result in partial or organ-specific abnormalities, such as midline liver or atrioventricular discordance, observed in our cohort.

The distinction between PCD-associated and non-PCD-associated HTX may not represent separate biological entities but rather a spectrum of ciliary and developmental abnormalities*. MNS1* provides an example of a gene that may influence laterality without causing classical PCD. Pathogenic variants in *MNS1* have been associated with situs abnormalities and infertility (25,27). Although MNS1 localises to respiratory cilia and sperm flagella and some outer dynein arm abnormalities have been reported, respiratory ciliary motility is largely preserved, consistent with a mild or absent respiratory phenotype. These findings support the hypothesis that some genetic defects may have different effects on nodal and respiratory cilia. Subtle defects may be sufficient to disrupt embryonic left–right patterning while remaining below the threshold required to cause classical respiratory PCD.

Respiratory symptoms typically associated with PCD were less frequent in the non-PCD group. However, the absence of differences for some symptoms likely reflects referral bias, as patients with recurrent respiratory infections are more likely to undergo PCD assessment. Nevertheless, some HTX patients without PCD displayed mild respiratory phenotypes despite respiratory ciliary structure and function not meeting current PCD diagnostic criteria (19). Subtle ciliary abnormalities may therefore have limited impact on respiratory clearance but greater consequences during embryonic left–right patterning. These findings should not be interpreted as representative of the general HTX population, as this cohort was recruited from a specialist PCD diagnostic service within a tertiary respiratory centre.

Specific laterality patterns differed between HTX groups. Midline liver was more common in HTX patients without PCD, and several other abnormalities, including malrotation, biliary atresia, and gastrointestinal atresia, were also more frequent. These findings may indicate a distinct laterality phenotype in some non-PCD HTX patients. Biliary atresia has been associated with syndromic laterality defects and congenital cholangiociliopathies, further supporting potential links between ciliary biology and organ development (28–30).

Cardiovascular defects also differed between groups, with atrioventricular discordance significantly more common in HTX patients without PCD. This may represent a subgroup of HTX associated with genetic pathways distinct from PCD. Larger multicentre studies incorporating whole genome sequencing will be required to identify potential causative variants, particularly as new candidate laterality genes continue to emerge.

The heterogeneous nature of HTX represents a challenge when defining patient cohorts, as laterality abnormalities exist across a clinical spectrum. We therefore used a broad clinical definition of HTX to capture the range of presentations encountered in practice. This approach allowed comparison of patients with and without PCD and may help distinguish classical motile cilia disorders from alternative mechanisms affecting left–right patterning.

The ultrastructural defects identified in HTX-PCD patients were consistent with previous studies, including absence of outer dynein arms, combined outer and inner dynein arm defects, and nexin-dynein regulatory complex abnormalities (31–33). No central pair or radial spoke defects were identified, consistent with the absence of central pair structures in embryonic nodal cilia and their limited role in situs determination (32,34). Some patients had normal TEM findings, which is compatible with *DNAH11*-associated PCD, where defects may require three-dimensional TEM or immunofluorescence for detection (35).

In contrast, TEM analysis of HTX patients without PCD identified a subset with extra microtubules or microtubular disorganisation affecting >5% of cilia. These abnormalities are usually considered secondary changes associated with infection or inflammation and are not recognised PCD defects. However, analysis of >2,800 individuals undergoing PCD assessment demonstrated that these ultrastructural abnormalities were associated with congenital heart disease and, for extra microtubules, with dextrocardia. Their lower frequency in PCD patients further supports that they do not represent classical PCD. Instead, these findings raise the possibility that subtle axonemal abnormalities, previously considered secondary changes, may reflect defects affecting nodal cilia function and contribute to laterality abnormalities.

Quantitative HSVM analysis further supported the presence of subtle functional differences in HTX patients without PCD. Previous studies have reported ciliary dysmotility in HTX patients, with a mean prevalence of approximately 46.2% (Table 5). In our cohort, PCD patients demonstrated reduced beat parameters compared with controls, whereas HTX patients without PCD generally showed preserved or increased ciliary beating amplitude.

**Table 5.** Previous studies on CHD in HTX patients.

| Study (Author, Year) | Population (number of patients) | Patients with cilia dysmotility |
| --- | --- | --- |
| Nakhleh et al., 2012 (8) | HTX (43) | 40.0% |
| Garrod et al., 2014 (9) | 218 with CHD (39 with HTX) | 51.8% (61.0%) |
| Harden et al., 2014 (10) | CHD-HTX (14) | 100.0%<sup>a</sup> |
| Zahid et al., 2018 (13) | TGA (75) | 57.0% |
| Stewart et al., 2018 (15) | non-HTX CHD (63) | 32.0% |
| Zhao et al., 2024 (16) | CHD-HTX (69) | 41.0% |
| This study | HTX (43) | 32.6% |

<sup>a</sup> Enrolled only CHD HTX patients with cilia dyskinesia abnormalities Average of the previous studies list above excluding *Harden et al., 2014* is 46.2%

Within the non-PCD group, patients with microtubular disorganisation had higher beat amplitude compared with controls and patients without clear ultrastructural abnormalities. These findings suggest that alterations in axonemal structure can influence ciliary function even in the absence of diagnostic PCD. Although these changes may have limited effects on respiratory clearance, they could contribute to abnormal nodal cilia function during embryonic development. Further multicentre studies integrating genetic, structural, and functional approaches are required to define the contribution of subtle ciliary abnormalities to HTX.

## Data Availability

Data is included within the manuscript and supporting information. Further data that support the findings of this study can be made available on reasonable request.

## Footnotes

LV and MB are joint first authors.

## Author Contributions

Conceptualisation: LV, AS, TB. Samples processing and imaging: LV, MB, RR, RM, GB, EH, MD, SJI, DJMR, TB. Image and video assessment: LV, MB, TB. Data compilation and statistical analysis: LV, MB, TB. Supervision: TB. Manuscript – original draft: LV, MB, TB. Writing – Review & Editing: all authors.

## Patient and public involvement

Patients and/or the public were not involved in the design, or conduct, or reporting, or dissemination plans of this research.

## Ethics statements

Patient consent for publication: Not applicable.

This study includes retrospective data from subjects were referred for PCD diagnostic testing at the Royal Brompton Hospital between 1997 and 2024 and was approved by the Guy’s and St Thomas NHS Foundation Trust Electronic Record Research Interface (GERRI) under IRAS ID: 257283 and Rec Reference: 20/EM/0112.

## Acknowledgments

We would like to thank Guy’s and St Thomas’ NHS Foundation Trust Electronic Record Research Interface (GERRI) for their support throughout this project. Study authors and data contributors participate in the BEAT-PCD clinical research collaboration, supported by the European Respiratory Society.

## AI use statement

During the preparation of this work the author(s) used ChatGPT in order to assist with drafting, language refinement, and clarity of expression. After using this tool/service, the author(s) reviewed and edited the content as needed and take(s) full responsibility for the content of the published article.

