## Supplementary data for "Respiratory ciliary abnormalities in heterotaxy beyond primary ciliary dyskinesia: functional and ultrastructural characterisation"

**Respiratory cilia abnormalities in patients with heterotaxy: a functional and ultrastructural characterisation**

### Supplementary data

**Table S1. p-values comparing Non-HTX Non-PCD patients against HTX-PCD and HTX-only.** See Table 1 for PCD-related respiratory phenotype and diagnostic results at the time of the referral.

| | Phenotype | *p*-value (Non-HTX, Non-PCD vs HTX-PCD) | *p*-value (Non-HTX, Non-PCD vs HTX-only) |
|---|---|---|---|
| **Clinical features** | Median age (years) | 0.6940[#] | 0.1817[#] |
| | Sex (Male) | 0.2908* | 0.1324* |
| | Median nNO level (nL/min) | 0.0703[#] | 0.7857[#] |
| **Respiratory Phenotype** | Persistent wet cough | **<0.0001*** | 0.2502* |
| | Persistent rhinitis | **<0.0001*** | >0.9999* |
| | Episodes of otitis media | >0.9999* | 0.5435* |
| | Recurrent chest infections | 0.3604* | >0.9999* |
| | Hearing defects, glue ear | **0.0222*** | >0.9999* |
| | Term neonatal respiratory distress | **0.0029*** | 0.2303* |
| **Cilia motility** | Normal motility | **0.0114*** | **0.0006*** |
| | Immotile | **<0.0001*** | >0.9999* |
| | Reduced beat amplitude | >0.9999* | 0.2890* |
| | Mixed beat pattern | **0.0030*** | **0.0059*** |
| **Cilia Structure[§]** | Reported PCD ciliary ultrastructural defects | **<0.0001*** | >0.9999* |

[#] Mann-Whitney test
* Fisher's exact test
[§] Results that included the assessment of less than 50 cilia were excluded (due to occasional cross-section obliqueness, dynein arms cannot be assessed as readily as other ciliary structures)
[†] These included 8 cases with outer dynein arm defects, 1 case with defects of inner dynein arms and 6 cases with defects of both dynein arms
p-values in bold indicate statistical significance ($p<0.05$)

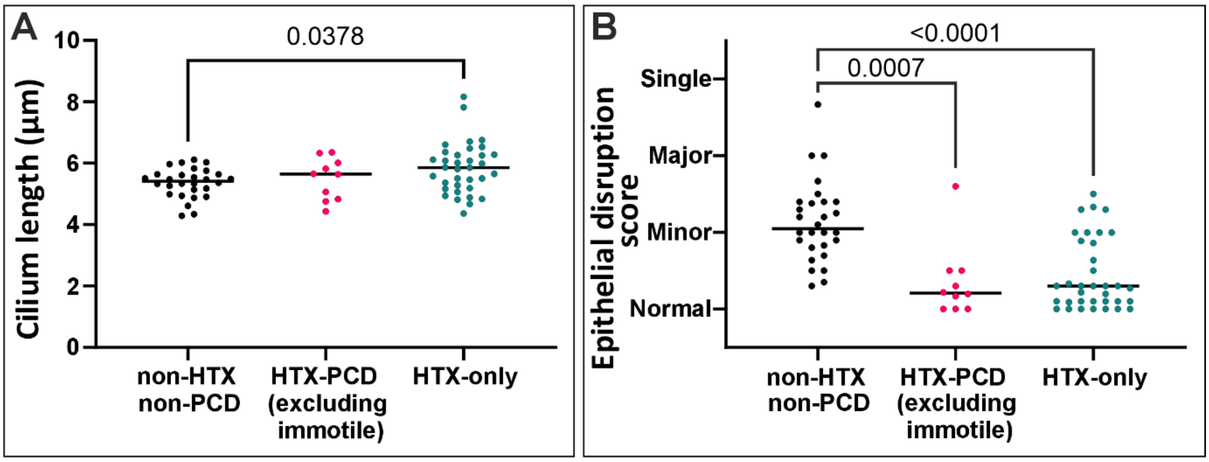


**Supplementary figure 1. Further parameter measured from diagnostic high-speed video-microscopy data of airway cilia from control (non-HTX, non-PCD) and HTX patient with or without PCD**. (A) The HTX-only subjects had a greater cilium length compared with the control group. (B) Both the HTX-PCD and HTX only subjects had videos with less epithelial disruption compared with the control group. Statistical significance was determined using the Kruskal-Wallis test.

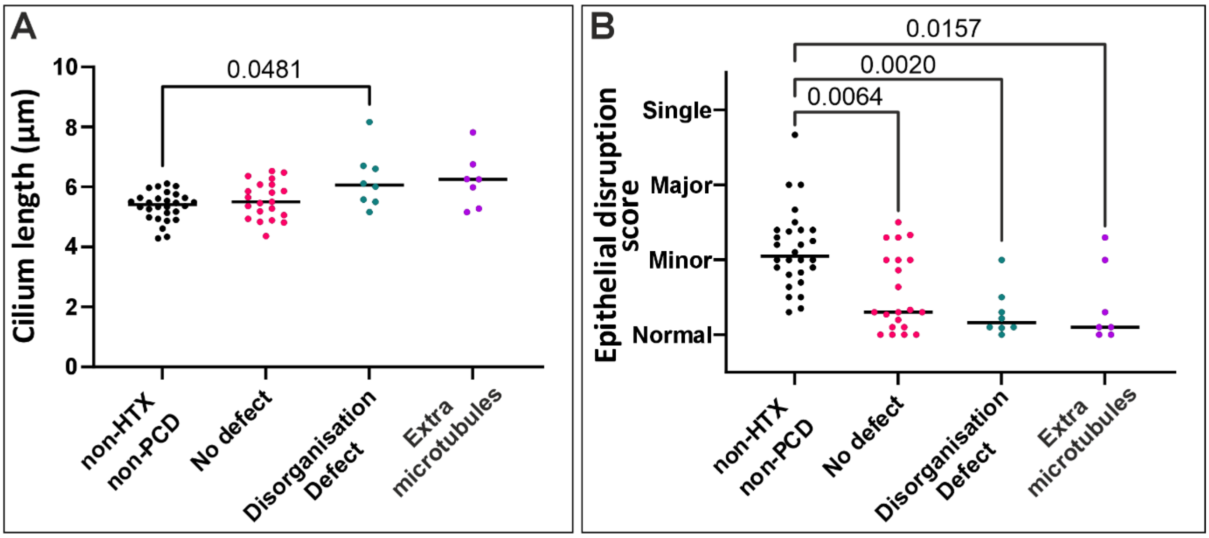


**Supplementary figure 2. Further parameter measured from diagnostic high speed video microscopy data of airway cilia from control (non-HTX, non-PCD) and HTX -only that have been grouped on the presence of ultrastructural defects**. (A) The HTX-only subjects with a ciliary disorganised defect (>5% of their total ultrastructural defects recorded by transmission electron microscopy) had a greater a greater cilium length compared with the control group. (B) The three HTX-only epithelial groups, no defect (<5% ultrastructural defects), ciliary disorganisation defect (>5% ultrastructural defects) and extra microtubules (>5% ultrastructural defects) had less epithelial disruption compared with the control group. Statistical significance was determined using the Kruskal-Wallis test.
